# On an optimal testing strategy for workplace settings operating during the COVID-19 pandemic

**DOI:** 10.1101/2020.12.22.20248752

**Authors:** X. Hernandez, S. Valentinotti

## Abstract

High quality daily testing for the presence of the SARS-CoV-2 in workplace settings has become part of the standard and mandatory protection measures implemented widely in response to the current pandemic. Such tests are often limited to a small fraction of the attending personnel due to cost considerations, limited availability and processing capabilities and the often cumbersome requirements of the test itself. A maximally efficient use of such an important and frequently scarce resource is clearly required. We here present an optimal testing strategy which minimises the presence of pre-symptomatic and asymptomatic members of the population, derived under a series of simplifying assumptions, which however retain many of the generalities of the problem and yield robust results, as verified through a number of numerical simulations. We show that reduction in overall infected-person-days, *IPD*, by significant percentages can be achieved, for fixed numbers of tests per day of 5% and 10% of the population, of 30% and 50% in the *IPD* numbers, respectively.

## Introduction

Within the context of the present COVID-19 pandemic, it has become clear that thus far, the most efficient strategy towards reducing the spread of the disease includes strict social distancing rules, reinforcing basic hygiene measures and the imposition of lockdown policies on the part of local and national governments. This last however, must clearly be tempered by the obvious need to keep essential workplace facilities operating, with examples including hospitals, energy production, food production and distribution, pharmaceutical industries, etc. to mention but a handful of the most obvious such cases.

Continual operation of such facilities has firstly included the adoption of safety and hygiene protocols during working days, and crucially, strict Sanitary Checkpoints, protocols for the daily entrance of persons attending. Sanitary Checkpoints (SC) serve the purpose of identifying symptomatic individuals which can then be tested directly for the virus, or in any case, sent home for a certain safety period.

Unfortunately, in the present pandemic, SC implementation is a measure of only limited efficacy, due to the important contribution of pre-symptomatic and asymptomatic infected persons (1, 2). It is now clear that a substantial fraction of transmissions is in fact the result of interactions with pre-symptomatic and asymptomatic individuals (3). This has led to the implementation of testing strategies, where a certain sample of persons attending are selected for direct, accurate testing, typically through PCR or more recently antigen testing. These accurate tests however, were initially not widely available and remain relatively scarce in certain areas, particularly outside big cities and in developing countries (4) and are somewhat cumbersome to implement, as well as expensive and generally not amiable to massive, daily implementation. For the above reasons, in most facilities where PCR testing is used, only a small fraction of the regular work force is sampled on any particular day. Thus, we have a situation where it is of the utmost urgency to apply optimal strategies to select the daily test sample, a scarce and expensive resource that must clearly be used to maximal efficiency.

It is important to note that the identification of asymptomatic persons is not only of relevance for the safe operation of an essential production facility, but also as a clear means of reducing the overall extent of the pandemic, as the more infected individuals are identified anywhere (and hence identified for surveillance, contact tracing, early treatment and/or quarantine protocols), the more successful societies will be in controlling epidemics.

While the importance of small screening intervals has been pointed out (5) such approaches assume a copious availability of daily tests, a situation which is not always practicable. In this paper we present an optimal sample selection strategy which maximises the number of asymptomatic infected persons identified from a fixed population, under the restriction of a given fixed number of daily tests available. In Section 2 we present simple probabilistic arguments showing that a randomly selected daily sample, from which persons which have already tested negative within the immediate *τ*_*I*_ day period have been excluded, should minimise the total number of infected-person-days (henceforth *IPD*) over any fixed period of time, where *τ*_*I*_ is the timescale over which the general population of infected persons is replaced by a new one, under the common assumption of a constant 15 day infection period, *τ*_*I*_ = 15 days. In Section 3 we implement a number of numerical simulations following idealised populations subject to a number of infection probabilities, percentages and evolution of pre-symptomatic and asymptomatic fractions and fraction of available daily tests. We show that indeed the optimal strategy presented in Section 2 results in a minimal number of *IPD* over any fixed period of time, and that this result is general to the epidemiological evolution, a constant or rising overall infection fraction in the population.

While a number of idealised assumptions remain in the model, it can be shown that the conclusion is sufficiently robust to warrant attention as a further element in aiding the control of the present pandemic.

### Section 2. Probabilistic arguments

We begin with simple considerations, imagine a constant infection fraction phase of the epidemic at a particular location, where the average fraction of infected individuals, *I*_*f*_, is a small constant number. If the infection period for all, symptomatic or asymptomatic persons, is a constant *τ*_*I*_ number of days, and if we imagine a cohort of simultaneously infected persons, it is clear that over the period over which they are infected, they must each infect one healthy individual, to ensure *I*_*f*_ remains a constant. Hence, assuming the probability of becoming infected in any single day for a healthy person in the general population is a constant, *P*_*I*_, and taking a linear approximation for the cumulative infection probability, the probability of becoming infected after a *τ*_*I*_ day period, *P_τ_*, will satisfy:

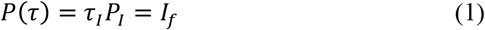

such that after a *τ*_*I*_ day period, each healthy individual has a chance *I*_*f*_ of having been infected, and therefore, the next cohort of infected individuals again represents the same fixed *I*_*f*_ fraction of the total. If the fraction of infected individuals is a constant *I*_*f*_ it is reasonable to assume that on average, the probability that a healthy individual has of becoming infected on any particular day, will be a constant, *P*_*I*_. From the above linear approximation, given values of *I* _*f*_ and *τ*_*I*_, the daily infection probability can be estimated as:

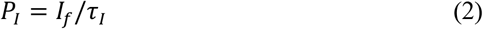

It is clear that cohorts of infected individuals will not be temporally exclusive, but will occur in a scrambled fashion over time, not altering the above equation, provided *I*_*f*_ and *τ*_*I*_ remain constant over time. If additionally, a certain person had a negative test result on a particular day, we shall assume that the probability of that individual being infected that day is zero, *i*.*e*. no false negatives are assumed on tests being performed. Thus, the probability of that individual becoming infected the following day becomes *P*_*I*_, and the probability of the individual not becoming infected on the day following his negative test is (1 − *P*_*I*_). The probability of not becoming infected after two days following the negative test is now (1 − *P*_*I*_)^2^, and in general, on average, after *n*, days, the probability of being infected becomes a non-linear function given by:

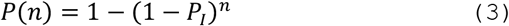

which corrects the linear approximation given previously. Again, over a *τ*_*I*_ day period, the above probability must yield *I*_*f*_, if this fraction is to remain constant, so that:

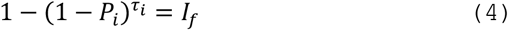

From this equation *P*_*I*_ can be calculated, for given values of *I*_*f*_, and *τ*_*I*_. Notice that for *P*_*I*_ ≪ 1, a Taylor expansion of equation 4 to first order recovers the zero-order intuitive result of equation 1. In order to develop an optimal testing strategy, we shall assume that we have a fixed number of persons attending the facility in question every day, having, in the absence of any test, an equal infection probability as the general population, *I*_*f*_. Thus, the first order testing strategy, if *N*_*T*_ is the number of tests available every day, is simply to draw a random sample of *N*_*T*_ members from the attending population at the workplace setting every day. However, notice that if someone tested negative on one particular day, the probability that this person is infected will begin to grow from zero on the days following the negative test, while the probability that a random member of the population is infected, prior to any tests, will be *I*_*f*_. Thus, we should endeavour to test attending persons having the highest probability of being infected, so that the limited number of tests available are used optimally towards identifying more effectively the infected members. Clearly, a member who has tested negative on a particular day should be excluded from the testing pool until the probability of this member being infected becomes again equal to that of the average population. Such a member should be excluded from the testing pool for a period *n*_*ex*_ such that:

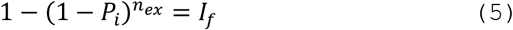

Clearly, equations 4 and 5 are identical, and therefore,

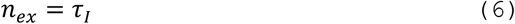

which is the main result of this section. A graphic representation of this is presented in Figure 1, which shows the logarithm of the probability of being infected after an *n* day period, *P*(*n*) of equation (3), for *P*_*I*_ = 6.69798 × 10^−4^, corresponding to a steady infection fraction of *I*_*f*_ = 0.01 for *τ*_*I*_ = 15.

**Figure 1:**
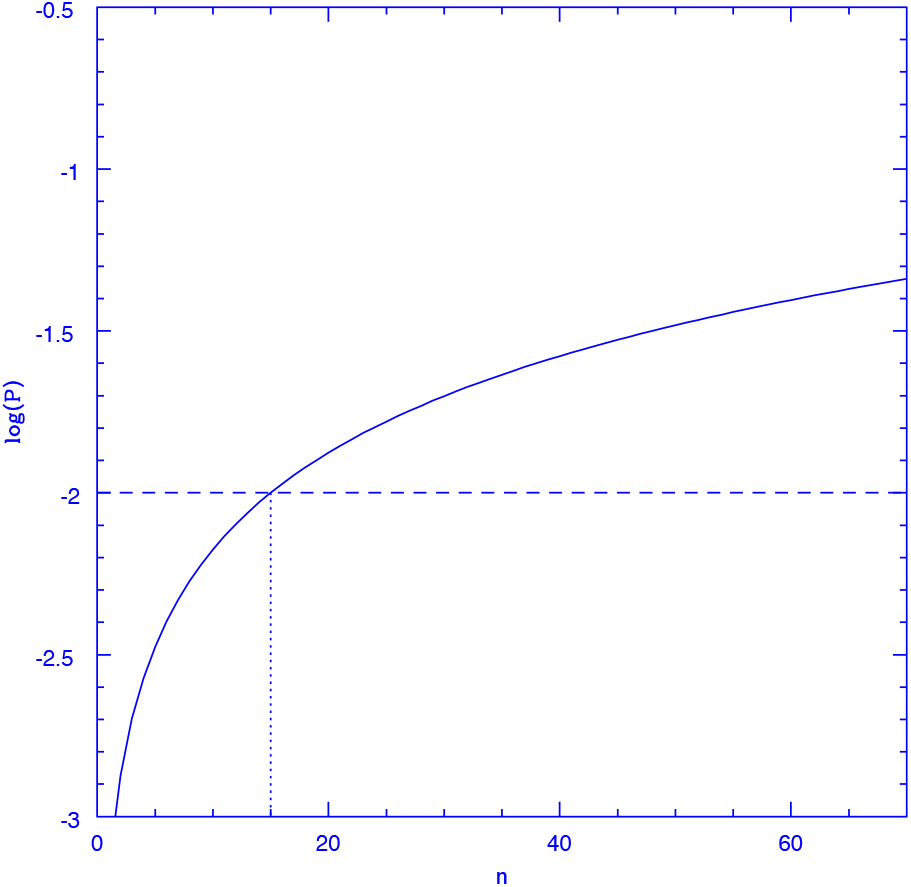
The figure shows the probability of an individual which tested negative on day zero of being infected on subsequent days, for *P*_*I*_ = 6.69798 × 10^−4^, corresponding to a steady infection fraction of *I*_*f*_ = 0.01 and *τ*_*I*_ = 15. This probability remains below that of the average population, 0.01, for all days before *n* = 15, after which, it remains above this number indefinitely.

As can be seen from the figure, the probability of being infected grows from zero on the day of the negative test, gradually with *n*, and converges to a value of 1 as *n* tends to infinity. The horizontal line shows the value of *I*_*f*_ = 0.01, the average probability of being infected for the general population. Clearly, *P*(*n*) overtakes *I* _*f*_ precisely at *n* = *τ*_*I*_ = 15 days.

Currently, the 15 most affected countries report close to 1/1000 COVID-19 deaths per capita (6), which assuming a 0.5% infection fatality rate yields 20% as the fraction of the population which has been infected in these countries, on average. Assuming further a 10-month current duration for the pandemic and a 15-day infection period, yields an average infected fraction of 1% for these countries. We hence take 1% for this variable, as a broad average reference value. Although approximately constant values of *I*_*f*_ over time have been observed for the current pandemic on a variety of places and times, e.g. the slowly rising bursts seen in some countries, or the extended troughs between waves often seen in others (6), in general, *I*_*f*_ will be a function of time. While to the accuracy presently available *τ*_*I*_ appears to be close to a constant, a situation where *I*_*f*_ = *I*_*f*_(*t*) will be more common, leading also to *P*_*I*_ = *P*_*I*_(*t*). Interestingly, in such a case, the factors (1 − *P*_*I*_)^*x*^ in equations 4 and 5 will both be replaced by the same function of time,

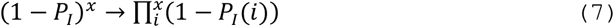

Clearly, again, the optimal testing strategy remains unchanged, with *n*_*ex*_ = *τ*_*I*_ This conclusion applies whenever *P*_*I*_ is a slowly varying function of time (in comparison to *τ*_*I*_, any sudden spikes, e.g.the influx of a considerable fraction of infected persons added to the total population, will clearly invalidate the argument above). It is of course possible that a member testing negative on a particular day becomes infected on the following one. If such case happens to be one of the asymptomatic ones, he will be allowed to attend while infected for the 15-day period over which he is infected, and presumably infectious. This however is less likely than that an average member who has not been tested might be infected, as can be seen from Figure 1. With a limited number of tests per day, one can at best minimise the number of infected persons attending, but driving this number to zero is unfortunately impossible, unless the daily testing of everyone might become an option.

Finally, we consider the effects of a having a finite total sample. It is clear that as time goes on, for certain fractions of the total population being tested, it could well happen that on a certain day there are not enough members who have not been tested within the previous 15 days to complete the *N*_*T*_ tests available that day. Given the positive definitive character of the probability function shown in Figure 1, for any value of its parameters, it follows that the optimal sampling strategy becomes to exclude from the daily testing pool any members who have been tested within the previous 15 days, while if that rule cannot be met without reducing *N*_*T*_, the sample should be augmented by adding all members who have gone 14 days without testing. If this again does not allow to complete the *N*_*T*_ tests available, all those who have gone 13 days without testing are considered, and so on.

Notice that once finite population effects begin to appear and the available tests have to be made up by including members not having been tested over periods smaller than *τ*_*I*_, the testing sample becomes drawn from a mixed population, including members who have not been tested over a range of recent days. Hence, taking *n*_*ex*_ values in this range (which depends on the fraction of tests available and the epidemiological details, as that will determine the average rate at which infected persons are found and sent home, which in turn affects the numbers remaining, from which the daily sample is to be drawn) will result in a softening of the resulting *IPD*(*n*_*ex*_) curves, the expected infinite population minimum at *n*_*ex*_ = *τ*_*I*_ can become broad, with a complex dependence on the details.

### Section 3. Numerical simulations

We now describe and show the results of a number of numerical simulations following sample populations under a variety of assumptions, where we can assess the generality of the scheme presented. We model a population of 1,000 members evolving under the following rules: Initially all members are assigned a healthy or infected status with a probability (1 – *I*_*f*_) or *I*_*f*_, respectively, with *I*_*f*_ = 0.01. Of those assigned as infected, a random sample of 25% are assigned as displaying symptoms and the rest as not displaying any symptoms, these last are a mix of the pre-symptomatic and the asymptomatic ones. During every following day, the epidemiological evolution considers a probability of becoming infected of *P*_*I*_ = 6.69798 × 10^−4^ for each healthy member. Again, 25% of those newly infected are assigned as displaying symptoms at the onset. Also, all those infected and not displaying symptoms are assigned, only on their 5th infected day, a 60% probability of passing from infected and not displaying symptoms to infected and displaying symptoms. This fixes a final asymptomatic fraction of 0.4 × 0.75 = 0.3, in accordance with recent estimates (2). Further, any infected members having spent more than 14 days in this state are returned to a healthy status. This ensures a steady *I*_*f*_ = 0.01 value, on average.

Then the intervention is imposed, any members displaying symptoms are sent home for a 14-day period, under the assumption of an efficient SC. Then, a testing sample as described in the previous section is constructed, which will have a variable number of *N*_S_ members from which a fixed *N*_*T*_ members are selected for testing at random every day. Figure 2 shows results for *N*_*T*_ = 50 and *N*_*T*_ = 100, left and right panels respectively. This is done for a fixed *n*_*ex*_ value, and the simulation is run for 200 days. Every day the number of infected persons attending is recorded to determine the IPD resulting. Given the stochastic nature of the problem, the whole 200-day simulation is repeated 10,000 times to gauge the intrinsic variance present. This is then repeated for various values of *n*_*ex*_ in the range shown in Figure 2. The solid lines give the average over the 10,000 realisations of the *IPD* values obtained over the second 100-day period. The first 100-day period is run to allow for a steady state to develop. The dashed lines give the 1*σ* intervals for the *IPD* values shown, over the 10,000 realisations, which for the parameters modelled are sufficient to reach convergence, further increase in the number of realisations considered yields only marginal differences from the results reported.

**Figure 2:**
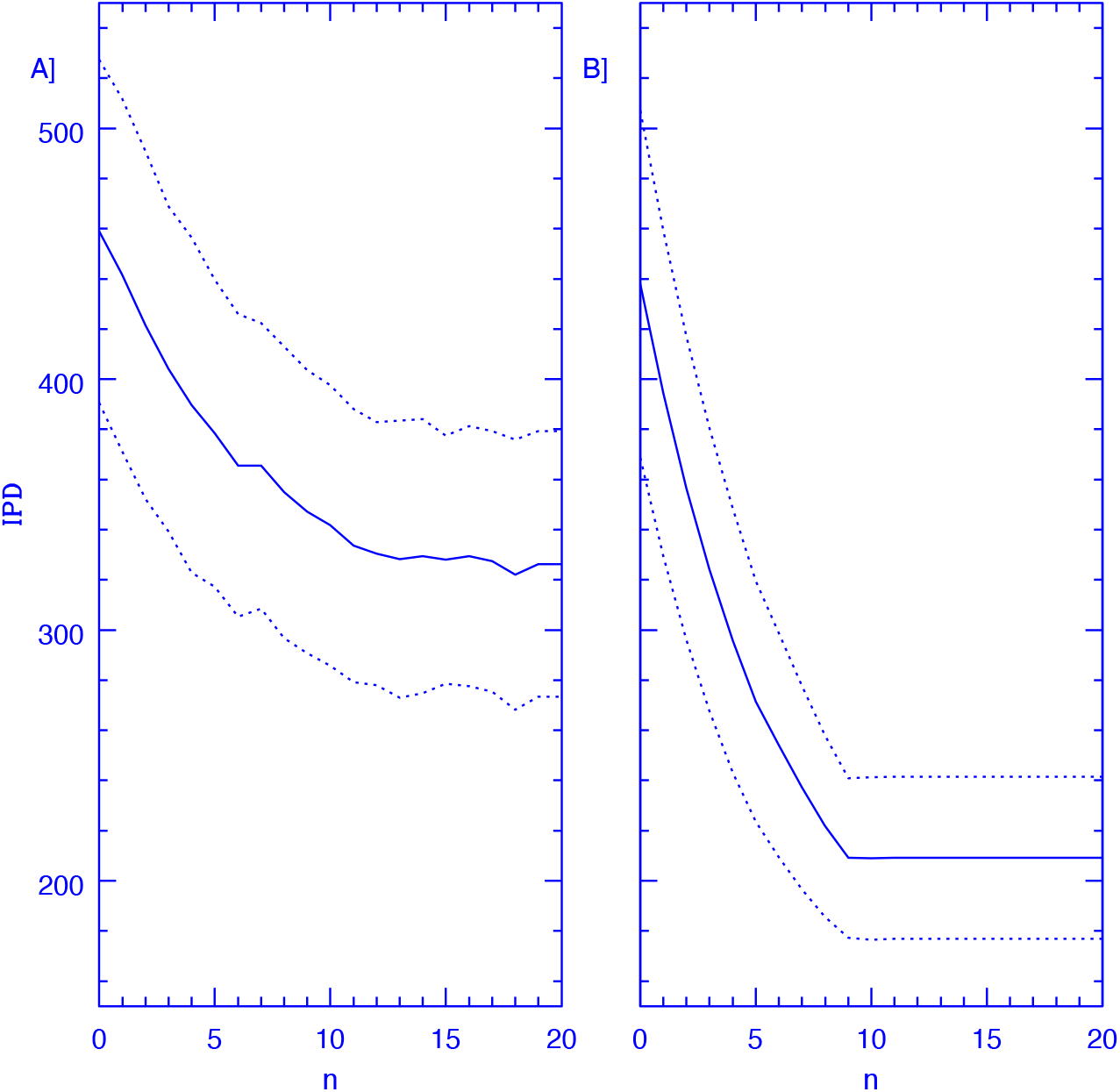
The figure shows IPD values over a 100-day period, red curves, obtained for a population of 1000 members attending a facility where 50 (A panel) and 100 (B panel) PCR tests are performed every day on a random sample of members, from which persons which test negative on a particular day are excluded for the subsequent *n* days, as a function of *n*. Members testing positive are sent home for a 14 day period. The epidemiological model corresponds to a value of *I*_*f*_ = 0.01 which remains constant over time. The solid lines show the stochastic variance inherent to the problem, giving 1σ intervals over 10,000 realisations, this last corresponding to the dashed line. An efficient SC is also assumed.

In absence of any intervention, one expects an average value of *IPD* = 1,000 × *I*_*f*_ × 100 = 1,000 indeed, we get 1000 ± 118 for this quantity over the 10,000 realisations. The introduction of the SC alone reduces these values to 484 ± 73, a reduction of a little more than a factor of two. If one then also includes a random sampling of 50 tests from which none of the members attending are excluded, we obtain an *IPD* = 458 ± 68, as shown by the *n*_*ex*_ = 0 point in the left panel of Figure 2. We see that the pure random sample achieves only a small further reduction in *IPD* numbers, as persons recently tested (and hence having a very small probability of being infected) have an equal chance of being selected for testing as persons not having been tested over a longer period. As the number of days after test which result in exclusion from testing is increased, a clear drop in *IPD* appears. Then, optimising further by the careful construction of the testing sample described, reduces the *IPD* values still further, down to 322 ± 54 in the broad minimum around *n*_*ex*_ = 15. Hence, a very significant reduction of 30% in *IPD* numbers, at constant number of daily tests performed, is achieved merely by excluding recently tested persons from the daily samples.

In the right panel of Figure 2 we see that as the testing sample grows to *N*_*T*_ = 100, the optimisation through the exclusion of recently tested members yields much more important results, with the drop from *n*_*ex*_ = 0 to *n*_*ex*_ = 15 now being of a very sizeable 53% of the *IPD* numbers found at *n*_*ex*_ = 0, showing the power of the sampling strategy presented. In this case however, we see clearly the finite sample effects appearing in the convergence at *n*_*ex*_ = 9.

Beyond this point (for the 10% of the total population being tested daily) it becomes impossible to find a daily sample not including persons having not been tested over a *n* > 9 day period, and the method saturates. Notice that for *N*_*T*_ = 50 and *n*_*ex*_ > 10 the total *IPD* numbers become even smaller than what is obtained for the double number of daily tests but *n*_*ex*_ = 0.

Finally, Figure 3 is equivalent to Figure 2, but gives results for simulations where the underlying epidemiological model is one where *P*_*I*_ rises over time with a constant doubling timescale of 50 days, being equal to that of the case summarised in Figure 2 at 100 days. In this case, in the absence of any intervention we obtain *IPD* = 2077 ± 154. The introduction of an efficient SC reduces these numbers to *IPD* = 1329 ± 115. Again, the reduction achieved by the optimisation of the testing sample is important in both cases, when *N*_*T*_ = 50, of 28.5% of the *n*_*ex*_ = 0 values. Yet, in going to *N*_*T*_ = 100, shown in the right panel of Figure 3, the corresponding reduction is now of a much more significant 52%, achieved without increasing the number of tests performed, merely through an optimised testing strategy. Again, saturation through finite sample effects becoming apparent at *n*_*ex*_ = 9. As in the previous case, the convergence of the *N*_*T*_ = 50 simulation for large *n*_*ex*_ values actually occurs at *IPD* values smaller than what is obtained in the *N*_*T*_ = 100 one at *n*_*ex*_ = 0. Also illustrative of the strength of the method presented is that the fractional reduction in total *IPD* numbers shown, is in all cases stronger in going from *n*_*ex*_ = 0 to larger values, than what results from going from the pure SC case to the *N*_*T*_ = 50 and *N*_*T*_ = 100 ones at *n*_*ex*_ = 0.

**Figure 3:**
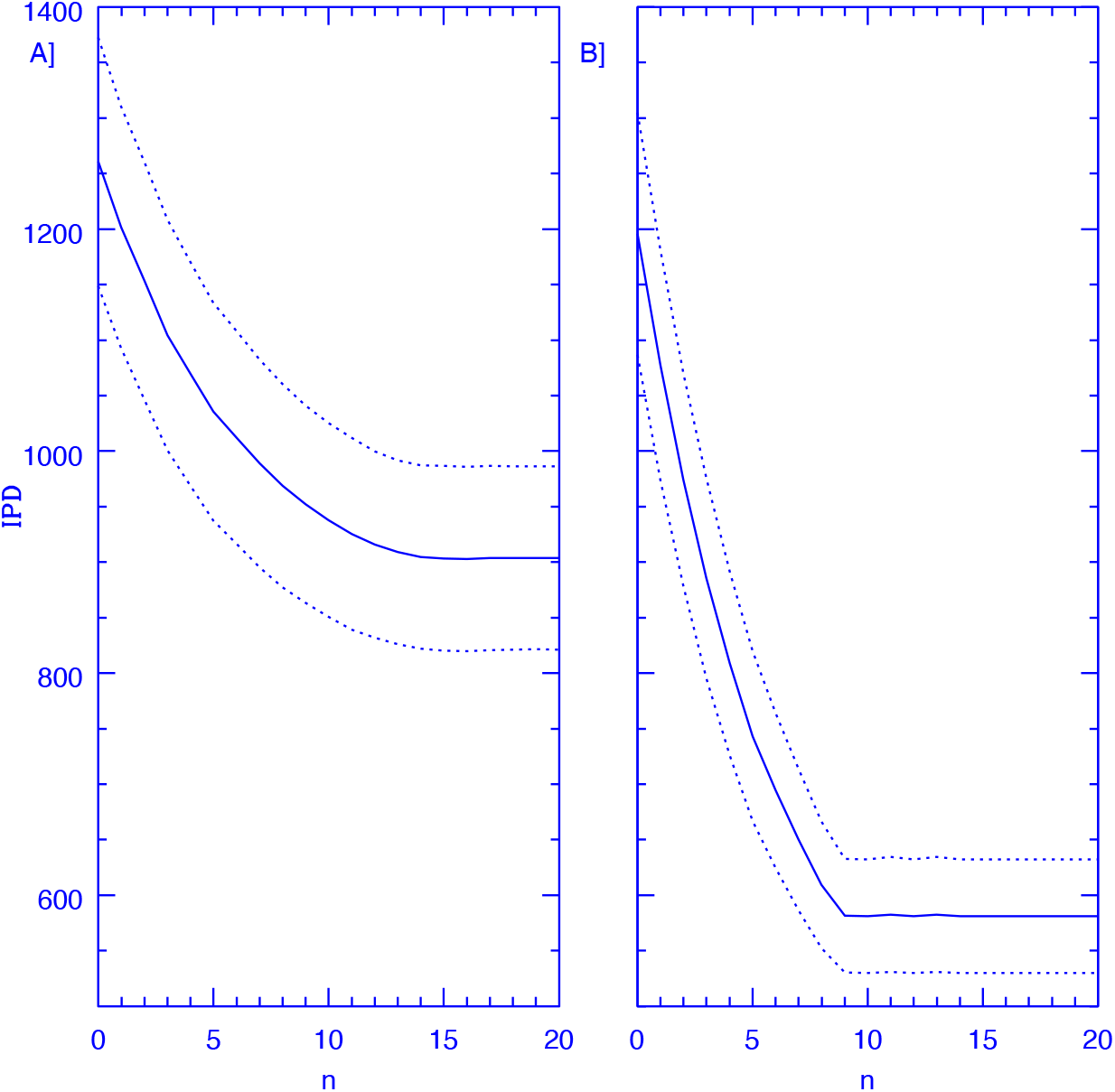
The figure shows IPD values over a 100-day period, red curves, obtained for a population of 1000 members attending a facility where 50 (A panel) and 100 (B panel) PCR tests are performed every day on a random sample of members, from which persons which test negative on a particular day are excluded for the subsequent *n* days, as a function of *n*. Members testing positive are sent home for a 14-day period. The epidemiological model corresponds to a value of *P*_*I*_(*t*) which grows linearly having a doubling timescale of 50 days and starting at *P*_*I*_ = 6.69798 × 10^−4^. The solid lines show the stochastic variance inherent to the problem, giving 1σ intervals over 10,000 realisations, this last shown by a dashed line. An efficient SC is also assumed.

A fairly general feature is that the reduction in IPD numbers obtained is fractionally smaller as *n*_*ex*_ increases, this is a direct consequence of the second derivative of the probability function of equation 5 being negative, as shown in Figure 1, the convergence of this probability function as *n* grows means that the largest drops in *IPD* numbers will come from the initial increases in *n*_*ex*_ away from zero, although the broad minimum remains at *n*_*ex*_ = 15, this last, provided no finite sample convergence appears.

The examples shown above are arbitrary and clearly extremely over simplified, not intended to represent any particular real situation. They do however serve to illustrate the usefulness of the scheme presented, as well as its generality and robustness to changes in the underlying epidemiological model. The actual reduction in IPD numbers will be sensitive to the details of the particular problem, but under a wide range of parameters, the conclusion of a daily random sample, from which persons that have been tested within the previous τ_*I*_ days were removed, as representing the optimal solution, will remain.

Further, we have included the assumption of 100% sensitivity in the PCR tests, while in reality this appears to grow rapidly from zero with time since infection to reach a maximum at around 8 days followed by an initially slow drop with time e.g. see the appendix in (5) and references therein. Therefore, the conclusion presented will not be modified, as re-testing within the first few days will not only be unlikely to detect an infected person (their chances of being infected being still small) but also, as these tests will be largely “wasted” through their sensitivity being low. The effect of the steeper decline in test sensitivity after about 13 days will be negligible in cases where the daily test percentage of the population is larger than 10%, where the saturation mentioned above appears before this period. We have taken an asymptomatic fraction towards the lower range of inferred values (2), with the intention of showing clearly the potential of the method presented, under less than optimal conditions for it; clearly, the effect of the method will tend to zero as the asymptomatic fraction goes to zero, and increase substantially as this fraction increases.

The scalings resulting from changes in the total population, as expected from basic probabilistic considerations, are for average resulting *IPD* values which remain constant as a fraction of the total population considered, with the corresponding 1σ intervals scaling with the square root of the total population considered, all other parameters being equal.

Fatalities are not explicitly included in the model, but will not have any significant effects provided the total fatality rate is low and/or any fatalities are promptly replaced from the general population. Finally, we note that the method presented can obviously be used in conjunction with pooling strategies, which, whenever sample taking logistics and infection prevalence allow (see for example (7)), permit increases in the total number of daily tests.

## Section 4. Discussion

We have presented an optimal testing strategy designed for minimising the number of infected asymptomatic persons present during a pandemic in a workspace setting, under the constraint of a fixed number of tests per day. The infected population substitution timescale, which corresponds to the infected period for the simple case where all infected individuals remain infected/infectious for exactly the same constant period of time, τ_*I*_ is identified as the critical parameter of such an optimisation. This raises two important points i) The optimal use of a finite number of high-quality tests is to perform a random sampling of the present population from which members having been tested during the preceding τ_*I*_ days are excluded, and which is augmented by including those members not having been tested during the preceding τ_*I*_ − 1 days, and so on, until the fixed number of tests per day can be randomly chosen, *i*.*e*. until the testing sample is larger than the number of daily tests. ii) It is crucial to have a more accurate and population weighted estimate of τ_*I*_, which surely is only a universal constant to a very first approximation.

## Data Availability

No data not publicly available has been used.

## Section 5. Acknowledgements

X.H. acknowledges support from Universidad Nacional Autónoma de México DGAPA grant IN-106220 and Consejo Nacional de Ciencia y Tecnología CONACyT.

